# Association of genetic risk and physical activity with incident type 2 diabetes

**DOI:** 10.1101/2025.02.14.25322281

**Authors:** Xuan Zhou, Germán D. Carrasquilla, Malene R. Christiansen, Roelof A.J. Smit, Lars Ängquist, Torben Hansen, Ruth J.F. Loos, Niels Grarup, Allan Linneberg, Tuomas O. Kilpeläinen, Jordi Merino

## Abstract

**Objective:** To assess whether daily step counts and genetic risk interact to influence the risk of developing type 2 diabetes.

**Research Design and Methods:** We analyzed data from 9,501 participants in the *All of Us* Research Program with both genetic and wearable device–derived physical activity data and without diabetes at baseline. Physical activity was quantified using daily step counts. Genetic risk was assessed using a global polygenic score. Incident type 2 diabetes was identified using electronic health record–linked diagnostic codes. Multivariable Cox proportional hazards models estimated hazard ratios (HRs) for type 2 diabetes across genetic risk and physical activity levels. We tested for additive interaction using the relative excess risk due to interaction (RERI). In secondary analyses we used physical-activity intensity measures using wearable-derived and self-reported intensity levels.

**Results:** Type 2 diabetes incidence rates ranged from 4.1 per 1,000 person-years (95% CI, 2.5–5.7) in individuals with high physical activity and low genetic risk to 18.4 (95% CI, 15.2–21.6) in those with low physical activity and high genetic risk (HR, 6.2 (95% CI: 3.97, 9.6)). A significant additive interaction was observed (RERI, 0.20; 95% CI, 0.04–0.36; P = .007), with 15% (95% CI, 2–27) of excess risk attributed to the interaction. Similar interaction patterns were found using device-based intensity metrics and self-reported physical activity measures.

**Conclusions:** These findings provide evidence of additive interactions between genetic risk and physical activity, underscoring the potential value of integrating genomic and device-derived data to identify individuals who would more likely benefit from increasing physical activity.

**Graphical abstract:** 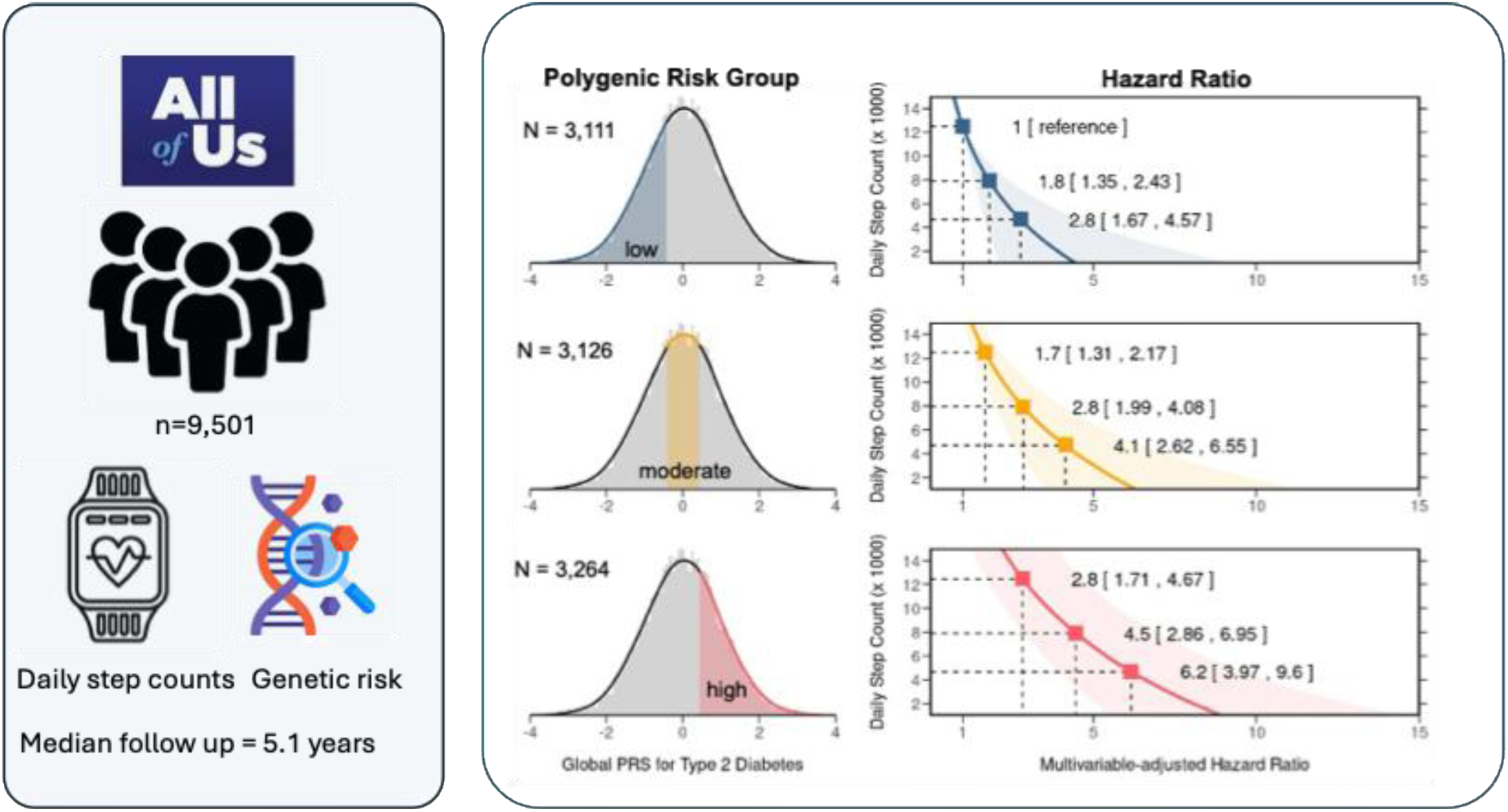

**Article Highlights:** 

**Why did we undertake this study?:** Despite longstanding interest in gene–environment interactions influencing the development of type 2 diabetes, identifying such interactions in human populations remains challenging, partially due to difficulties in accurately measuring environmental exposures. Step count is an objective, interpretable measure of daily ambulatory physical activity that offers a novel opportunity to evaluate these interactions. To date, no studies have specifically investigated how daily step counts may interact with genetic susceptibility to influence type 2 diabetes risk.

**What is the specific question(s) we wanted to answer?:** To what extent do genetic risk and physical activity, measured via daily step count, independently and jointly contribute to the risk of developing type 2 diabetes?

**What did we find?:** We observed a gradient of increasing type 2 diabetes risk associated with high genetic susceptibility and low step count. This gradient deviated from additivity, with the combination of high genetic risk and low physical activity accounting for substantial proportion of excess type 2 diabetes risk. The evidence of additive interactions between genetic risk and physical activity was replicated in an independent cohort using self-reported physical activity data and longer-term follow-up.

**What are the implications of our findings?:** Our results highlight the joint impact of genetic risk and physical inactivity on type 2 diabetes development. These findings support the integration of genetic and behavioral risk factors in risk stratification and prevention strategies and may inform targeted public health interventions aimed at reducing the global burden of type 2 diabetes.

## INTRODUCTION

The risk of developing type 2 diabetes varies considerably among individuals, reflecting differences in susceptibility to the often collectively called environmental factors (1,2). This variability underpins the significance of gene-environment interaction, wherein an individual’s genetic profile, together with its downstream molecular effectors, interact with environmental exposures to uniquely modulate disease risk (3–6). While gene-environment interactions are pervasive in nature, shaping phenotypes such as plant growth (7), bird migration (8), and grasshopper adaptation to new environments (9), identifying such interactions in the context of human traits and diseases remains challenging (1), with often conflicting findings in the literature (10–13). This challenge is partly due to the limited precision and depth of environmental exposure assessments, particularly when self-reported (14,15).

Step count provides a simple and interpretable metric of daily ambulatory activity, offering advantages over self-reported physical activity measures or less intuitive accelerometer-derived intensity classifications (16). Prior studies have shown that higher step counts are associated with lower type 2 diabetes risk (17,18), and that accelerometer-measured intensity-specific physical activity may attenuate type 2 diabetes genetic risk (19). However, previous studies of gene–physical activity interactions have generally relied on single time-point physical activity assessments and have not accounted for the dynamic nature of physical activity over time. Moreover, no prior study has directly investigated how daily step counts interact with genetic risk on the development of type 2 diabetes.

In this study, we analyzed longitudinal data from a population-based cohort in the United States, comprising 9,501 adults with objectively measured physical activity over time, to test the hypothesis that physical activity, measured via daily step count, interacts with genetic risk in the development of type 2 diabetes. We further evaluated whether similar patterns were observed using additional activity measures, including device-based intensity levels and self-reported physical activity.

## METHODS

### Study design and population

We used data collected from participants in the *All of Us* Research Program. Details on the design and execution of the *All of Us* have been published previously (20,21). In this study, we used Curated Data Repository (CDR) version 7 for Controlled Tier (C2022Q4R9). This dataset included information on physical measurements and vital signs collected at enrollment, surveys, electronic health record (EHR), Fitbit data, and genetic information from participants enrolled between May 6, 2018, and July 1, 2022. Our analyses focused on participants who owned a Fitbit and agreed to share their Fitbit, EHR, and genetic information. We excluded participants who did not wear a Fitbit for at least 6 months. Only the authorized authors who completed *All of Us* Research Program Responsible Conduct of Research Training accessed the deidentified data from the Researcher Workbench (a secured cloud-based platform). Since the authors were not directly involved with the participants, institutional review board review was exempted in compliance with *All of Us* Research Program policy.

In a secondary analysis, we used data from Inter99 (21). Inter99 is a population-based pre-randomized lifestyle intervention study aiming to prevent ischemic heart disease and type 2 diabetes. Baseline data were collected from March 1999 until January 2001 and there were 6,784 participants. Follow-up was conducted after one, three, five, ten, and twenty years for a health examination, completion of questionnaires and risk assessment.

### Physical activity assessment

Activity tracking data for this study came from the Bring Your Own Device program that allowed individuals who already owned a tracking device (Fitbit, Inc) to consent to link their activity data with other data in the *All of Us* cohort. By registering their personal device on the patient portal, participants could share all activity data collected since the creation of their personal device account. We used mean daily steps calculated on a monthly basis for each participant. Consistent with prior physical activity data curation approaches (17), days with less than 10 hours of wear time, less than 100 steps, or greater than 45 000 steps or for which the participant was younger than 18 years were removed. In addition, we removed months with fewer than 15 valid days of monitoring. To capture additional dimensions of physical activity, we derived device-based measures of intensity, including daily lightly active minutes, moderate active minutes, and very active minutes []. We also analyzed self-reported physical activity data from the Inter99 study, assessed via validated questionnaires covering work and leisure-time activity (22), at baseline and at one-, three-, and five-year follow-ups. Reported activity levels were converted into minutes per week by summing responses based on a five-day working week.

### Type 2 diabetes polygenic scores

We generated a global polygenic score for type 2 diabetes that captures overall genetic burden using genetic variants and weights identified in a previous study that did not include participants from this study (23). We followed standard procedures to filter genetic data to include only variants that had passed initial quality control (24). We used PRS–continuous shrinkage (PRScs) (25) to infer posterior SNP effect sizes under continuous shrinkage priors, with the scaling parameter auto-estimated (25). Scores were computed with ancestry-specific variants and weights.

### Ascertainment of type 2 diabetes

Incident type diabetes cases were identified using any incident billing code in EHR. We excluded any new diagnoses coded during the first six months of monitoring, assuming that such conditions were likely prevalent, but not yet recognized clinically. The EHR data from different participating sites were mapped and harmonized using the Observational Medical Outcomes Partnership common data model (26). In Inter99, we used data from separate nationwide health registries to define type 2 diabetes events.

### Statistical analysis

A flow diagram describes how many participants were excluded based on the criteria used to create the analytical dataset (Supplementary Figure 1). Descriptive statistics for participant’s demographic and clinical characteristics were presented by median and IQR for continuous variables and frequency for categorical variables.

We used Cox proportional hazard models to examine associations of genetic risk and step counts with type 2 diabetes risk. For these analyses, we censored cases that occurred 6 months after completing enrolment to mitigate potential bias due to time-varying confounding. Person-time for each participant was calculated from the six months after completing baseline assessment to the diagnosis of type 2 diabetes, death, loss to follow-up, or the end of the follow-up period, whichever came first. We modelled polygenic scores and step counts as continuous variables. We averaged daily step counts within each month and subsequently treated step counts as a time-varying covariate in the Cox models to account for month-to-month variations in physical activity during the follow-up period. We also classified participants according to categories of genetic risk and physical activity (nine categories based on thirds of genetic risk and step counts, with low genetic risk and high physical activity as reference) and conducted stratified analyses. We adjusted the multivariable models for age (in years, continuous), sex (male, female) and genetic ancestry inferred from the first 10 principal components of genomic relationship matrix. We verified the proportional hazards assumption of the Cox model by using the Schoenfeld residuals technique (p values for the global test > 0.1). Analyses were conducted for complete cases given the small number of people with missing covariates. In secondary analyses, we further adjusted our models for BMI (quintiles of kg/m^2^). The rationale for not including BMI in the main model was because BMI can act as a collider or a mediator (27).

We tested for additive and multiplicative interactions of genetic risk and physical activity with type 2 diabetes incidence. For additive interactions we assessed the relative excess risk due to interaction (RERI) and further examined the decomposition of the joint effect—that is, the population attributable proportions to genetic risk alone, physical activity alone, and to their interaction(28). For these analyses, we used the following formula (RERI = RR11 - RR10 - RR01 + 1) and (attributable proportion (AP) = RERI/ RR11) (28). CIs for each of the interaction measures were calculated using the delta method described by Hosmer and Lemeshow. We tested for multiplicative interactions using the log-likelihood ratio test to compare the goodness of fit of a multivariable-adjusted model with and without the cross-product interaction term(29). For these analyses, we considered genetic risk and physical activity as continuous variables and modeled the effect per 1SD.

In secondary interaction analyses, we derived device-based measures of physical activity intensity, including daily lightly active minutes, fairly active minutes, and very active minutes. These intensity levels were defined using metabolic equivalents (METs): 1.5–3.0 METs for lightly active, 3.0–6.0 METs for fairly active, and >6.0 METs for very active minutes (30,31). We also incorporated total self-reported physical activity data from the Inter99 study, categorizing participants as having high (7–12 h/week) or low (0–2 h/week) activity. Additional analyses were conducted stratifying participants by ethnic group (European, African, East Asian, and South Asian) and by sex.

Two-sided tests were used to assess whether polygenic risk for T2D and physical activity are associated with T2D incidence. A one-sided test was used to assess whether RERI is greater than 0, i.e., testing the hypothesis that the combined effects of polygenic risk for T2D and physical activity exceeds their individual effects. All statistical analyses were performed using R software, version 4.0.3 (R Foundation). We followed the Better Precision-data Reporting of Evidence from Clinical Intervention Studies & Epidemiology (BePRECISE) reporting guideline (32).

## RESULTS

Of 14,200 participants with available Fitbit data at the time of our analysis, 9,501 had valid physical activity data, genetic information, and no diabetes at baseline (Supplementary Figure 1). The median age was 56 years (interquartile range (IQR) 42–66), the median body mass index (BMI) was 28.3 kg/m^2^ (IQR 24.8–33.3), and most participants were women (72%), of European ancestry (88%), and held a college degree (72%; Table 1). Participants contributed 11.6 million person-days of Fitbit monitoring over a median of 5.7 years (IQR, 3.6–7.7), yielding 97.9 billion recorded steps. Median daily step count was 7,891 (IQR, 5,748–10,471), and step count showed a normal distribution (Supplementary Figure 2). Compared to non-include All of US participants, included individuals were more likely to be younger, female, of European ancestry, with lower BMI and higher educational attainment (Supplementary Table 1).

**Table 1.**
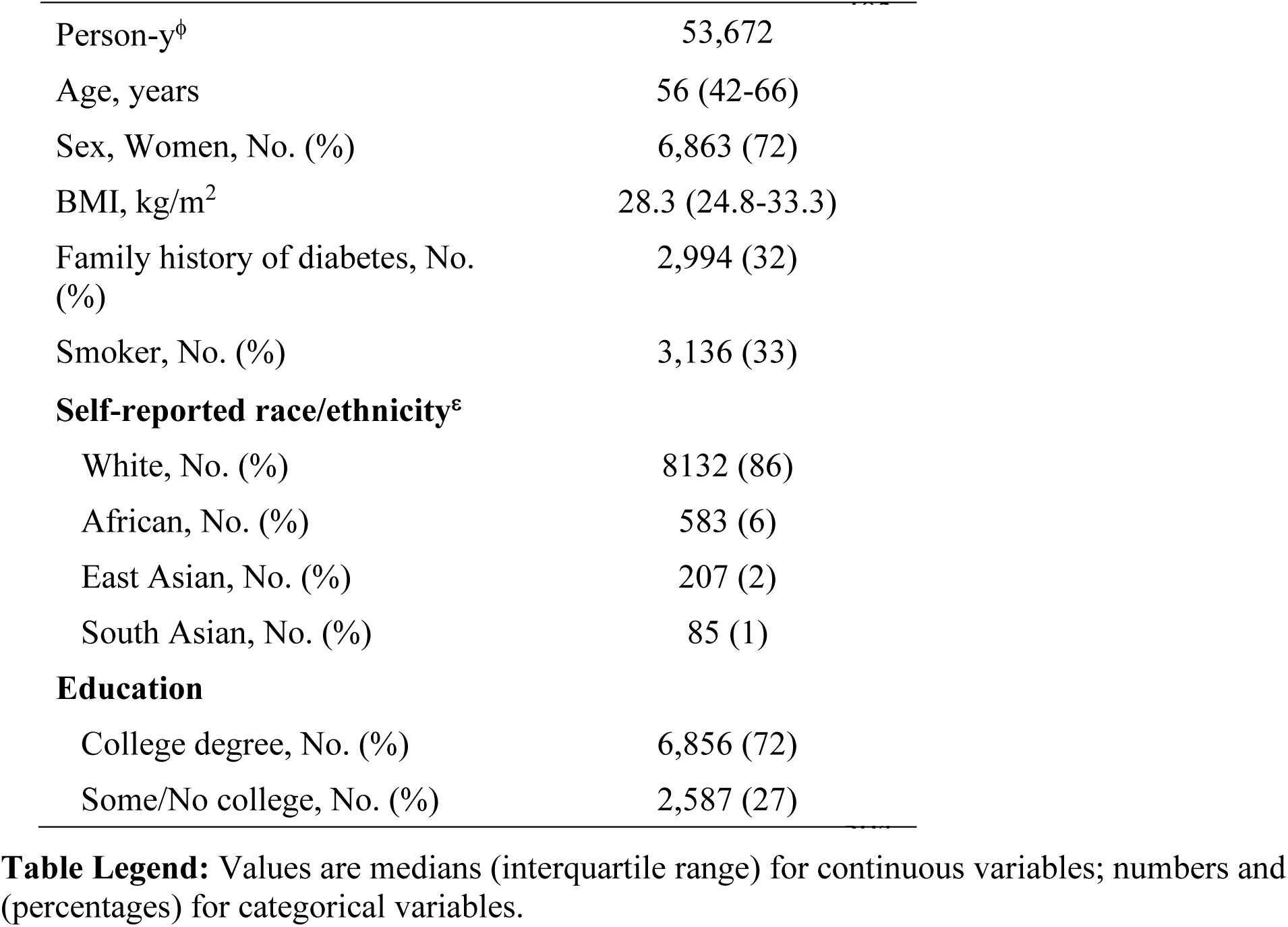
Baseline characteristics of the included *All of US* research program participants.

During 53,672 person-years of follow-up, 494 incident cases of type 2 diabetes were documented. Crude type 2 diabetes incidence rates per 1,000 person-year according to daily step counts tertiles were 13.5 (95% CI: 11.9-15.2), 7.5 (95% CI: 6.3-8.8), and 5.9 (95% CI: 4.7-7.0) for the lowest, intermediate, and highest tertile, respectively (multivariable-adjusted HR of 0.40 (95% CI: 0.26, 0.63; Table 2). Each standard deviation (SD) increase in step count was associated with lower risk of type 2 diabetes, with an adjusted HR of 0.65 (95% CI, 0.57–0.73).

**Table 2:** Adjusted hazard ratios of T2D risk according to tertiles of daily step counts and genetic risk.

|  | Low step count | Intermediate step count | High step count | P for trend |
| --- | --- | --- | --- | --- |
| No. of events/person-years | 263 / 19415 | 136 / 18,046 | 95 / 16,210 | — |
| Incidence rate (1,000 person-years; 95% CI) | 13.5 (11.9-15.2) | 7.5 (6.3-8.8) | 5.9 (4.7-7.0) | — |
| Multivariable-adjusted model | 1.00 (Ref) | 0.68 (0.57-0.82) | 0.40 (0.26-0.63) | <0.001 |

|  | Low genetic risk | Intermediate genetic risk | High genetic risk | P for trend |
| --- | --- | --- | --- | --- |
| No. of events/person-years | 107/ 18,180 | 142/ 17,991 | 245/ 17,500 | — |
| Incidence rate (1,000 person-years; 95% CI) | 5.9 (4.8-7) | 7.9 (6.6-9.2) | 14 (12.2-15.8) | — |
| Multivariable-adjusted model | 1.00 (Ref) | 1.59 (1.43-1.76) | 2.50 (2.04-3.06) | <0.001 |

**Table Legend:** Hazards ratios and 95% CI for T2D risk. Daily step counts and genetic risk were categorized in three categories based on the distribution of physical activity and polygenic score. Cox proportional hazards models were adjusted for age (in years, continuous), sex (male, female), and the first 10 principal components of the genomic relationship matrix.

Similarly, type 2 diabetes incidence increased across tertiles of polygenic risk, with incidence rates per 1,000 person-years of 5.9 (95% CI: 4.8, 7.0) in the lowest, 7.9 (95% CI: 6.6, 9.2) in the intermediate, and 14.0 (95% CI: 12.3, 15.8) in the highest tertile. In adjusted models, participants in the highest tertile had 2.50 times the risk of type 2 diabetes compared with those in the lowest (95% CI, 2.04–3.06; Table 2). When modelled in the continuous scale, each SD increase in polygenic score was associated with an adjusted HR of 1.52 (95% CI, 1.38–1.67).

When examining joint associations, crude incidence rates ranged from 4.1 (95% CI, 2.5–5.7) per 1,000 person-years among participants with low genetic risk and high physical activity to 18.4 (95% CI, 15.2–21.6) among those with high genetic risk and low physical activity (Figure 1). The corresponding multivariable-adjusted HR was 6.20 (95% CI, 3.97–9.60). Within the high genetic risk group, individuals with low activity (∼5,000 steps/day) had nearly double the incidence of those with high activity (∼13,000 steps/day; HR, 1.92; 95% CI, 1.36–2.69). The joint association was higher than the sum of the risk associated with each factor alone, with a relative excess risk due to interaction (RERI) of 0.20 (95% CI: 0.04, 0.36); p=0.007). Decomposition of excess type 2 diabetes risk attributed 40% to genetic risk (95% CI: 31, 49), 45% to low physical activity (95% CI: 29, 62), and 15% to their interaction (95% CI: 3, 27; Table 3). No evidence of multiplicative interaction was observed (p>0.05).

**Figure 1:**
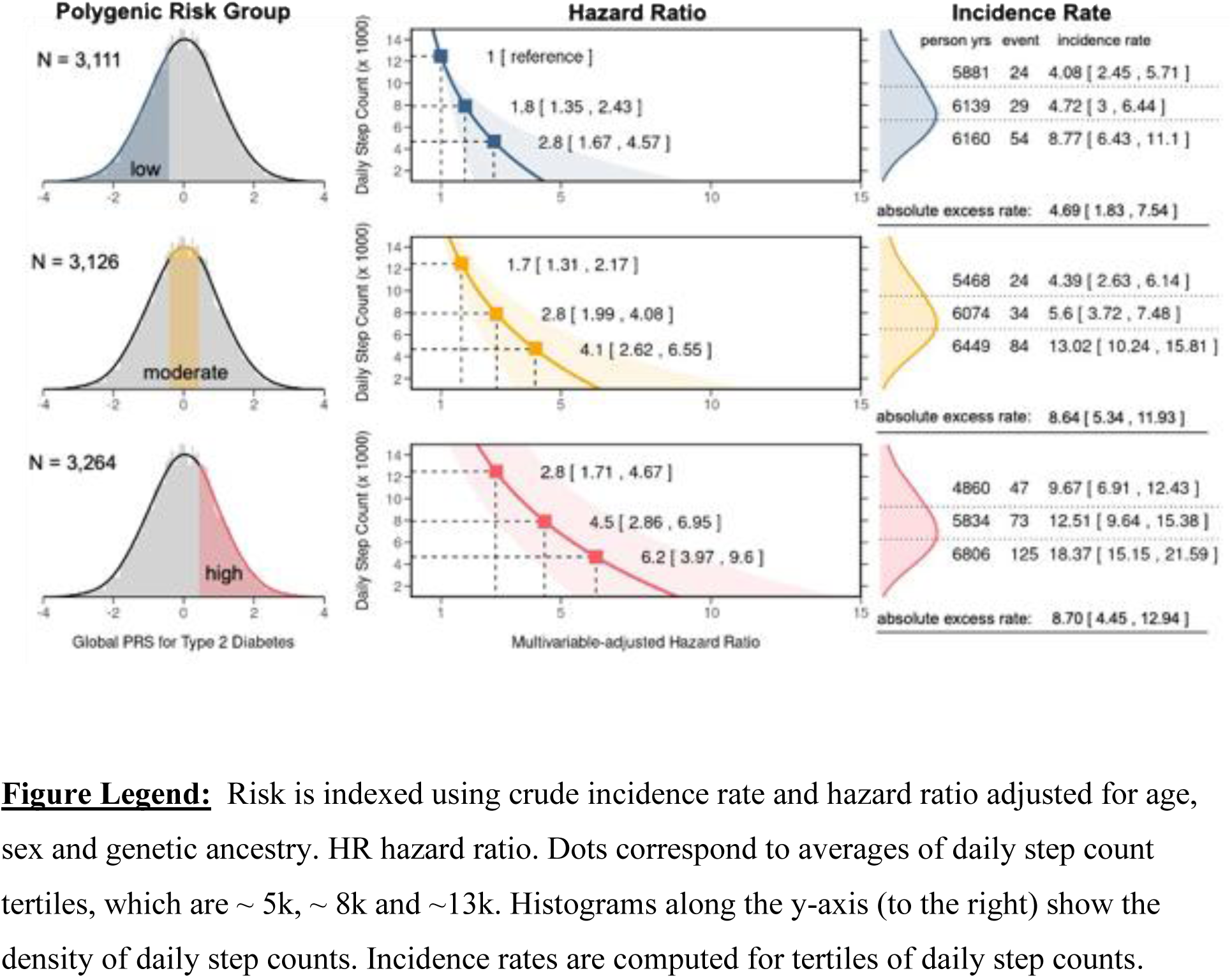
**Risk of incident type 2 diabetes by global polygenic risk score for type 2 diabetes, physical activity.**

**Table 3.** Additive interactions between physical activity and global polygenic risk of T2D.

|  |  |
| --- | --- |
| **Main effects** |  |
| Physical activity<sup>†</sup> | 1.53 (1.32, 1.74) |
| Polygenic score<sup>‡</sup> | 1.61 (1.32, 1.74) |
| Joint effect | 2.34 (1.87, 2.81) |
| **Relative excess risk due to interaction** |  |
| Relative excess risk due to interaction | 0.2 (0.04, 0.36) |
| *P* value | 0.007 |
| **Attributable risk proportion, %** |  |
| Daily step count | 39.8 (31.0, 48.5) |
| Polygenic score | 45.4 (28.6, 62.1) |
| Additive interaction | 14.8 (2.5, 27.2) |

**Table Legend:** Multivariable adjusted risk of type 2 diabetes estimated from Cox proportional hazards models adjusted for age, sex, and the first ten principal components of genomic relationship matrix. The joint analysis was conducted to quantitatively estimate the combined association of physical activity and genetic risk simultaneously with the incidence of type 2 diabetes. The relative excess risk due to interaction (RERI) is based on joint effects of a one-standard-deviation increase in PRS and a one-standard-deviation decrease in daily step count using the following formula ( $RERI = RR_{11} - RR_{10} - RR_{01} + 1$ ). The model's attributable risk proportion due to the interaction was calculated as $AP = RERI / RR_{11}(28)$ .
<sup>†</sup> per SD decrease in daily step counts.
<sup>‡</sup> per SD increase in the global polygenic scores.

We conducted a series of sensitivity and subgroup analyses. When physical activity was modeled by intensity, similar additive interactions were observed. The RERI was 0.21 (95% CI, 0.02–0.41; p=0.015) for moderate activity minutes, with 19% of excess risk attributable to interaction, and 0.40 (95% CI, 0.21–0.59; p<0.001) for highly active minutes, with 31% attributable to interaction (Supplementary Table 2). Adjustment for BMI did not materially change these estimates (Supplementary Table 2). Using total physical activity assessed by self-reported physical activity in Inter99, the RERI was 0.53 (95% CI, 0.30–0.76; p=0.007), with 44% of excess risk attributable to interaction (Supplementary Table 3). Adjustment for BMI attenuated this proportion to 38% (95% CI, –6 to 82; Supplementary Table 3).

In ancestry-stratified analyses, no significant associations between genetics and physical activity or interactions were observed in the African ancestry group (68 cases among 583 participants; Supplementary Table 4). Due to small sample sizes, models for South Asian (5 cases among 85 participants) and East Asian (9 cases among 207 participants) did not converge. Sex-stratified analyses showed additive interactions in both sexes. Among men, the RERI was 0.2 (95% CI: 0.01, 0.42; p=0.03), and 22% of excess risk was attributed to interaction. Among women, RERI was 0.2 (95% CI: -0.04, 0.43; p=0.050), with an attributable proportion of 11% (Supplementary Table 5).

## DISCUSION

Our findings provide consistent evidence of an additive interaction between genetic risk and physical activity in the development of type 2 diabetes. Specifically, individuals with both high genetic risk and low step counts had higher risk than expected from the sum of their individual effects, with approximately one-fifth of excess risk attributable to this interaction. Similar patterns were observed when physical activity was assessed using device-based intensity metrics or self-reported total physical activity. Since additive interactions reflect differences in absolute risk across populations (28), these observations may have potential public health relevance, suggesting that reducing the overlap of these risk factors could contribute to lowering type 2 diabetes incidence. However, replication in other cohorts and settings will be important to better understand the robustness and generalizability of these results.

Our study builds on prior research investigating gene-environment interactions in type 2 diabetes, which has often yielded inconsistent findings (10–13). One likely contributor to these discrepancies is error in exposure assessment due to reliance on self-reported methods (14,15). Using accelerometer-based physical activity measures, a recent UK Biobank study demonstrated an additive effect between physical activity and polygenic scores on T2D risk, with greater absolute risk reductions observed among individuals at high genetic risk who engaged in moderate-to-vigorous physical activity (19). Our findings extend this evidence base in three important ways. First, unlike the UK Biobank study, which relied on single time-point physical activity assessment, we modelled physical activity as a time-varying exposure, capturing behavioural changes over a median follow-up of 5 years. This approach more accurately reflects behavioral dynamics leading up to diabetes onset (33). Second, we used daily step counts as the primary exposure. Compared with intensity-based classifications such as “moderate-to-vigorous,” which may vary in interpretation and are less intuitive, step counts provide a simple, interpretable, and actionable metric for public health messaging (16). Third, our polygenic scores were derived from an external discovery dataset that excluded participants from this study, thereby mitigating potential inflation of type I error due to sample overlap, a methodological limitation that may have influenced the UK Biobank analyses. Together, our study strengthens the evidence that objectively measured physical activity interacts with genetic risk on the development of type 2 diabetes.

Physical activity remains central to public health recommendations for type 2 diabetes prevention (34), yet current guidelines do not account for heterogeneity in individual responses. Factors such as genetic susceptibility, body composition, and diet can all influence the effectiveness of physical activity (35). For example, in the ARIC cohort, the protective effects of physical activity were attenuated in women with higher genetic predisposition to insulin resistance (36), suggesting that greater physical activity may be needed to offset genetic risk. Similarly, a prior analysis from the All of Us Research Program estimated that individuals at high genetic risk for obesity would need to walk an additional ∼2,300 steps per day to match the obesity risk of those at lower risk (37). In our study, participants at high genetic risk who walked ∼5,000 steps per day had nearly double the incidence of type 2 diabetes compared to those walking ∼13,000 steps per day. Our models estimated that 15% of type 2 diabetes cases could have been prevented by eliminating one of the two risk factors, emphasizing the potential impact of increasing activity levels, particularly among genetically susceptible individuals.

Although the proportion of risk attributable to gene–physical activity interactions was similar when using step counts or device-based intensity measures, higher estimates were observed in the Inter99 cohort for total physical activity. This discrepancy may reflect differences in follow-up time, population characteristics such as age and obesity prevalence, or potential inflation due to self-reported activity measures. Since population-attributable risk depends on both exposure prevalence and effect size (38), exposure misclassification, especially with self-reported physical activity, could partly explain these differences. While these excess risk estimates due to the interaction may change over time with shifting diabetes incidence, our findings underscore the ongoing relevance of gene–physical activity interactions in type 2 diabetes.

This study has several limitations. First, as an observational study, participants were not randomized to high- or low-physical activity groups, limiting causal inference interpretations. Recall-by-genotype studies could help test these hypotheses under experimental conditions. Second, incident type 2 diabetes was defined solely based on any incident billing codes from the electronic health record, which may introduce misclassification. However, this approach is widely used in biobank research, including other All of Us analyses (17,39). Third, the number of type 2 diabetes cases was relatively small, reflecting limited follow-up and the subset of participants with linked wearable data. Fourth, while we used comprehensive polygenic scores for type 2 diabetes, other genetic risk factors (i.e., for obesity) were not included. Although we adjusted for BMI in secondary analyses, residual confounding, collider bias, or mediation effects could still exist. Larger studies with longer follow-up and detailed modelling of BMI trajectories may help address this. Finally, other molecular, clinical, or behavioral factors such as diet, body composition, or gut microbiome were not explicitly included in our analyses, but could affect risk or modify gene–environment interactions.

In conclusion, by quantifying the combined contributions of genetic risk and physical activity to type 2 diabetes risk, our results underscore the potential value of integrating genomic data with objective, device-derived measures of physical activity to better identify those who are most likely to benefit from targeted interventions. Future studies with larger, diverse populations and longer follow-up are needed to confirm these interactions and to evaluate how they can be translated into effective public health and clinical interventions.

## Data Availability

To ensure privacy of participants, data used for this study are available to approved researchers following registration, completion of ethics training and attestation of a data use agreement through the All of Us Research Workbench platform, which can be accessed via https://workbench.researchallofus.org/login. Code used for this study can be made available to users of the All of Us Research Workbench platform by contacting our study team.
Data from Inter99 are available upon request and approval.

## Funding/Acknowledgements

We gratefully acknowledge *All of Us* and Inter99 participants for their contributions, without whom this research would not have been possible. The *All of Us* Research Program is supported by grants 1 OT2 OD026549, 1 OT2 OD026554, 1 OT2 OD026557, 1 OT2 OD026556, 1 OT2 OD026550, 1 OT2 OD 026552, 1 OT2 OD026553, 1 OT2 OD026548, 1 OT2 OD026551, 1 OT2 OD026555, IAA AOD21037, AOD22003, AOD16037, and AOD21041 (regional medical centers); grant HHSN 263201600085U (federally qualified health centers); grant U2C OD023196 (data and research center); 1 U24 OD023121 (Biobank); U24 OD023176 (participant center); U24 OD023163 (participant technology systems center); grants 3 OT2 OD023205 and 3 OT2 OD023206 (communications and engagement); and grants 1 OT2 OD025277, 3 OT2 OD025315, 1 OT2 OD025337, and 1 OT2 OD025276 (community partners) from the National Institutes of Health (NIH).

This study was also supported by the Novo Nordisk Foundation grant NNF23SA0084103 and the European Union. JM was additionally supported by grants from the European Commission (HORIZON-EIC-2023-PATHFINDERCHALLENGES-01 - 101161509) and the EFSD/NNF Future Leaders Award (#0094134). Views and opinions expressed are however those of the author(s) only and do not necessarily reflect those of the European Union or European Innovation Council and SMEs Executive Agency (EISMEA). Neither the European Union nor the granting authority can be held responsible for them. RL was additionally supported by personal grants from the Novo Nordisk Foundation (Laureate award no. NNF20OC0059313) and the Danish National Research Fund (Chair DNRF161). TOK and MRC and were additionally supported by grants from the Novo Nordisk Foundation (NNF22OC0074128) and EFSD/Lilly European Diabetes Research Programme.

## Ethics declarations

The authors declare no competing interests. JM is the guarantor of this work, and, as such, had full access to all the data in the study and takes responsibility for the integrity of the data and the accuracy of the data analysis.

