## supplementary materials for "Association of genetic risk and physical activity with incident type 2 diabetes"

**Supplementary Figure 1.** Flow chart of study participants.

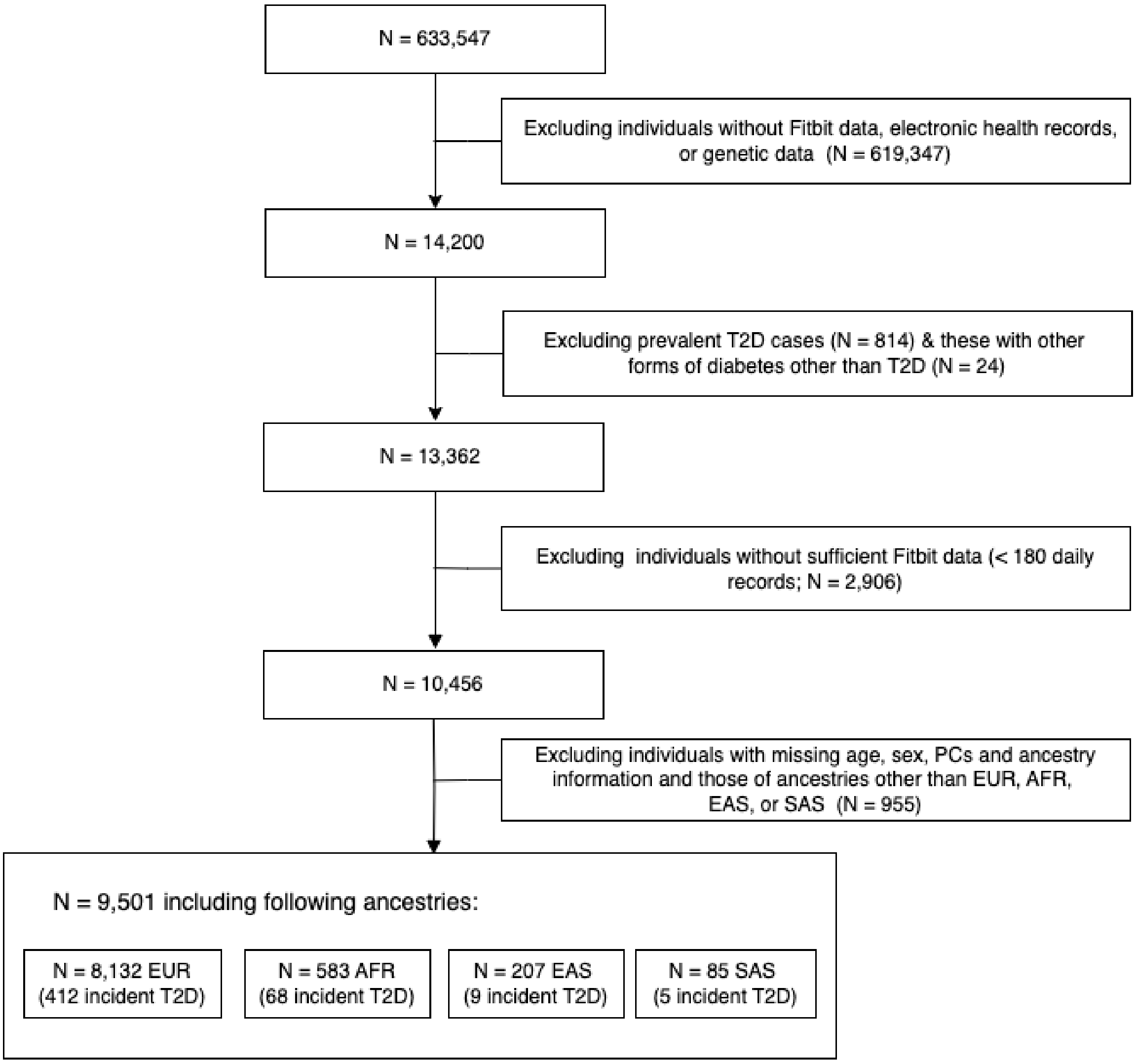


**Figure legend:** Among the 14,200 participants from the All of Us cohort with linked Fitbit data, electronic health records and genetic at the time of our analysis, 9,501 met the criteria for valid physical activity data at baseline and were at least 18 years old during the monitoring period. *EUR* European, *AFR* African, *EAS* East Asian, *SAS* South Asian.

**Supplementary Figure 2.** Distributions of daily step counts.

**A**

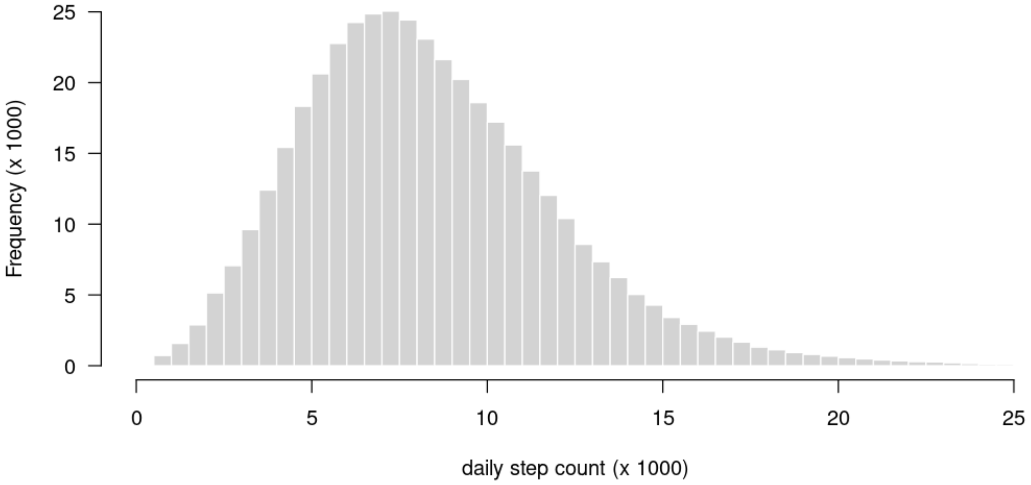


**Figure legend:** A. histogram of daily step counts in the *All of Us* cohort. Daily step counts followed an approximately normally distribution, with a median of 7,891 steps per day (IQR 5,748–10,471).

**Supplementary Table 1:** Baseline characteristics of participants included versus non-included in this study.

| | **Included (n=9,501)** | **Non-included (n=624,046)** |
|---|---|---|
| Age, years | 56 (42-66) | 52 (37-65) |
| Sex, Women, No. (%) | 6,863 (72) | 389,124 (63) |
| BMI, kg/$m^2$ | 28.3 (24.8-33.3) | 28.4 (24.4-33.6) |
| Smoker, No. (%) | 3,136 (34) | 219,829 (40) |
| **Self-reported race/ethnicity** | | |
| White, No. (%) | 8,230 (88) | 349,428 (68) |
| African, No. (%) | 582 (6) | 99,206 (19) |
| Others, No. (%) | 534 (6) | 66,128 (13) |
| **Education** | | |
| College degree, No. (%) | 6,856 (72) | 284,778 (46) |
| Some/No college, No. (%) | 2,587 (27) | 324,880 (52) |

**Table Legend:** Values are medians (interquartile range) for continuous variables; numbers and (percentages) for categorical variables.

**Supplementary 2 Table:** Additive interactions between global polygenic risk of T2D and physical activity by intensity

| | Lightly active minutes | | Moderate active minutes | | Very active minutes | |
|---|---|---|---|---|---|---|
| | *Model 1* | *Model 2* | *Model 1* | *Model 2* | *Model 1* | *Model 2* |
| **Main effect** | | | | | | |
| Physical activity† | 1.15 (0.97, 1.33) | 1.06 (0.92, 1.21) | 1.4 (1.14, 1.67) | 1.28 (1.06, 1.49) | 1.51 (1.26, 1.76) | 1.32 (1.12, 1.52) |
| Polygenic score‡ | 1.61 (1.34, 1.88) | 1.5 (1.24, 1.75) | 1.5 (1.19, 1.81) | 1.4 (1.13, 1.67) | 1.32 (0.96, 1.68) | 1.28 (0.95, 1.61) |
| Joint effect | 1.77 (1.42, 2.11) | 1.56 (1.27, 1.84) | 2.12 (1.64, 2.59) | 1.84 (1.46, 2.22) | 2.2 (1.74, 2.67) | 1.85 (1.48, 2.23) |
| **Additive interaction** | | | | | | |
| RERI | 0.01 (-0.14, 0.16) | -0.01 (-0.14, 0.13) | 0.21 (0.02, 0.41) | 0.16 (0, 0.33) | 0.37 (0.18, 0.56) | 0.25 (0.08, 0.43) |
| *P* value | 0.46 | 0.48 | 0.015 | 0.027 | <0.001 | 0.003 |
| **Attributable risk proportion, %** | | | | | | |
| Physical activity | 19.61 (3.59, 35.64) | 11.5 (-9.7, 32.7) | 36.2 (24.2, 48.2) | 33.1 (18.4, 47.7) | 42.6 (31.5, 53.7) | 37.4 (23.8, 51.0) |
| Polygenic score | 79.41 (54.22, 104.6) | 89.25 (62.5, 116) | 44.7 (23.6, 65.7) | 47.6 (23.2, 72.1) | 26.6 (2.9, 50.4) | 33.1 (3.1, 63.1) |
| Additive interaction | 0.97 (-18.4, 20.34) | -0.75 (-24.4, 22.9) | 19.1 (1.7, 36.6) | 19.3 (-1.3, 40.0) | 30.8 (11.6, 50.0) | 29.5 (3.7, 55.3) |

**Table Legend:** Multivariable adjusted risk of type 2 diabetes estimated from Cox proportional hazards models adjusted for age, sex, and the first ten principal components of genomic relationship matrix. Model 2 was further adjusted for BMI. The joint analysis was conducted to quantitatively estimate the combined association of physical activity and genetic risk simultaneously with the incidence of type 2 diabetes. The relative excess risk due to interaction (RERI) is based on joint effects of a one-standard-deviation increase in PRS and a one-standard-deviation decrease in physical activity measure using the following formula (RERI = RR11 - RR10 - RR01 + 1). The model's attributable risk proportion due to the interaction was calculated as AP = RERI/ RR11

† per SD decrease of the physical activity measure.

‡ per SD increase in the global polygenic scores.

**Supplementary Table 3** Additive interactions between physical activity and global polygenic risk of T2D among participants from the Inter99 study

| | *Model 1* | *Model 2* |
|---|---|---|
| **Main effects** | | |
| Physical activity† | 1.90 (0.93, 2.86) | 1.06 (0.46, 1.66) |
| Polygenic score‡ | 2.02 (1.32, 2.73) | 1.96 (1.22, 2.69) |
| Joint effect | 4.45 (2.30, 6.61) | 2.65 (1.29, 4.02) |
| **Relative excess risk due to interaction** | | |
| Relative excess risk due to interaction | 1.53 (0.30, 2.76) | 0.63 (-0.27, 1.53) |
| *P* value | 0.007 | 0.08 |
| **Attributable risk proportion, %** | | |
| Daily step count | 26.0 (10.9, 41.1) | 3.8 (-29.9, 37.5) |
| Polygenic score | 29.7 (9.3, 50.0) | 57.9 (9.4, 106.3) |
| Additive interaction | 44.4 (22.1, 66.6) | 38.3 (-6.3, 82.9) |

**Table Legend:** Multivariable adjusted risk of type 2 diabetes estimated from Cox proportional hazards models adjusted for age, sex, and the first ten principal components of genomic relationship matrix. Model 2 was further adjusted for BMI. The joint analysis was conducted to quantitatively estimate the combined association of physical activity and genetic risk simultaneously with the incidence of type 2 diabetes. RERI is based on joint effects of a one standard-deviation increase in the polygenic score and a reduction in weekly physical activity from 7-12h to 0-2h using the following formula (RERI = RR11 - RR10 - RR01 + 1). The model's attributable risk proportion due to the interaction was calculated as AP = RERI/ RR11[20].

† per SD increase in daily step counts.

‡ per SD increase in the global polygenic scores.

**Supplementary Table 4** Adjusted hazard ratios of T2D risk according to categories of genetic risk and physical activity among participants of African Ancestry.

| | **Low genetic risk** | **Intermediate genetic risk** | **High genetic risk** | **P for trend** |
|---|---|---|---|---|
| No. of events/person-years | 23 | 22 | 23 | — |
| Incidence rate (1,000 person-years; 95% CI) | 19.5 (11.5-27.4) | 19.7 (11.5-27.9) | 20.3 (12-28.6) | — |
| Multivariable-adjusted model | 1.00 (Ref) | 1.09 (0.8,1.47) | 1.17 (0.67,2.06) | 0.59 |
| | **Low PA** | **Intermediate PA** | **High PA** | **P for trend** |
| No. of events/person-years | 16 | 22 | 30 | — |
| Incidence rate (1,000 person-years; 95% CI) | 24.1 (15.5-32.7) | 19.3 (11.2-27.4) | 15.3 (7.8-22.7) | — |
| Multivariable-adjusted model | 1.00 (Ref) | 0.88 (0.7,1.11) | 0.73 (0.41,1.3) | 0.29 |

**Table Legend:** Hazards ratios and 95% CI for T2D risk according to categories of genetic risk or physical activity levels in an ancestry specific analysis conducted in All of Us. Cox proportional hazard models were adjusted for age (in years, continuous), sex (male, female), and genetic ancestry inferred from the first ten principal components of the genomic relationship matrix. There was no evidence of significant interactions between genetic risk and physical activity on the risk of T2D, with a relative excess risk due to the interaction estimate of 0.02 (-0.44-0.48, p value = 0.5)

**Supplementary Table 5** Interactions between genetic risk and physical activity in sex-stratified analyses

| | Men (n=2,618) | Women (n=6,883) |
|---|---|---|
| **Main effects** | | |
| Physical activity[†] | 1.41 (1.09, 1.73) | 1.65 (1.37, 1.93) |
| Polygenic score[‡] | 1.33 (0.79, 1.87) | 1.86 (1.43, 2.3) |
| Joint effect | 1.94 (1.27, 2.61) | 2.71 (2.07, 3.35) |
| **Relative excess risk due to interaction** | | |
| Relative excess risk due to interaction | 0.2 (-0.01, 0.42) | 0.2 (-0.04, 0.43) |
| *P* value | 0.03 | 0.05 |
| **Attributable risk proportion, %** | | |
| Daily step count | 43.6 (23.3, 64) | 37.9 (29.1, 46.7) |
| Polygenic score | 34.9 (-5, 74.8) | 50.6 (33.5, 67.7) |
| Additive interaction | 21.5 (-7.8, 50.8) | 11.4 (-1.2, 24) |

**Table Legend:** Multivariable adjusted risk of type 2 diabetes in men and women separately estimated from Cox proportional hazards models adjusted for age and ancestry-derived principal components.

† per SD decrease in daily step counts.

‡ per SD increase in the global polygenic risk scores
